# Serum potassium elevation and acute respiratory acidosis during thoracoscopic esophagectomy with intrathoracic carbon dioxide insufflation: a multicenter retrospective observational study

**DOI:** 10.64898/2026.07.04.26357266

**Authors:** Sakura Okamoto, Tokiko Tochii, Ran Hotta, Kento Nakamura, Jyunya Nakada, Hideaki Note

## Abstract

**Background:** Thoracoscopic esophagectomy with intrathoracic carbon dioxide insufflation and lung collapse, usually performed in the prone position, can markedly alter respiratory physiology and acid-base balance. Serum potassium elevation is often observed during acute respiratory acidosis in these procedures, despite the conventional view that respiratory acidosis has little effect on potassium. We quantified the intraoperative potassium change and explored associated factors.

**Methods:** This multicenter retrospective study included adults undergoing thoracoscopic esophagectomy with carbon dioxide insufflation in the prone or lateral decubitus position during 2022–2024. Arterial blood gas variables were evaluated after anesthesia induction and at the time of the lowest arterial pH during carbon dioxide insufflation. The primary outcome was the paired difference in serum potassium. Sensitivity, subgroup, and regression analyses were performed.

**Results:** All 131 patients were included: 117 in the prone position and 14 in the lateral decubitus position. Serum potassium increased from 3.96 ± 0.38 to 4.59 ± 0.63 mEq/L (mean increase, 0.64 mEq/L; 95% confidence interval, 0.55–0.73; p < 0.001). During the same period, pH decreased from 7.387 to 7.247 and arterial carbon dioxide tension increased from 41.48 to 58.60 mmHg. After excluding marked metabolic acidosis, the increase remained significant and similar in magnitude (0.618 mEq/L). In the centered multivariable model, lactate change was independently associated with potassium change (β = 0.303; p = 0.004), whereas arterial carbon dioxide change and preoperative renal function were not. The intercept remained significantly positive.

**Conclusions:** A clinically meaningful potassium increase was observed during thoracoscopic esophagectomy with carbon dioxide insufflation, while acute respiratory acidosis developed during the same period. The increase persisted after excluding marked metabolic acidosis and may not be explained solely by metabolic stress. Arterial blood gas assessment, including potassium measurement, is warranted during significant hypercapnia, and potential electrolyte consequences should be considered when permissive hypercapnia is accepted.

## Introduction

Carbon dioxide (CO₂) insufflation during thoracoscopic surgery for esophageal cancer has become widely used to facilitate exposure of the operative field. The prone approach has been reported to be associated with fewer respiratory complications and less blood loss than the lateral decubitus approach [1]; however, it can also substantially affect ventilatory management. Partial lung collapse and intrathoracic CO₂ insufflation, both of which are commonly used during these procedures, may cause marked changes in respiratory physiology and acid–base balance.

Clinically, elevations in serum potassium are often observed during these operations in association with acute respiratory acidosis. Although acidosis is known to influence potassium distribution [2], respiratory acidosis has traditionally been considered to cause little or no increase in serum potassium, compared with metabolic acidosis [3]. Recent clinical observations, however, have challenged this conventional view in critically ill patients. For example, Pathak and Nanda reported cases of severe, refractory hyperkalemia in patients with COVID-19 in which acute respiratory acidosis was considered a major contributing factor [4]. Nevertheless, it remains unclear how the combination of thoracoscopic CO₂ insufflation and ventilatory restriction modifies the expected physiological response.

The primary aim of this study was to quantify the magnitude of serum potassium elevation associated with acute respiratory acidosis during thoracoscopic esophagectomy performed with CO₂ insufflation in the prone or lateral decubitus position. We also sought to explore the physiological factors underlying this phenomenon, including the relative contributions of respiratory and metabolic components. Ultimately, this study aimed to contribute to safer ventilatory strategies and more appropriate electrolyte management during these complex procedures.

## Methods

### Study design

This was a retrospective, two-center observational study conducted at Aichi Medical University Hospital and Aichi Cancer Center. The study was conducted according to a previously published protocol [5], with modifications reflecting final confirmation of the surgical position and actual analytical procedures.

The published protocol primarily focused on prone-position thoracoscopic esophagectomy. However, during final data confirmation, some eligible cases at one institution were found to have been performed in the lateral decubitus position under thoracoscopic CO₂ insufflation. Therefore, the final study population was defined as adult patients undergoing thoracoscopic esophagectomy with intrathoracic CO₂ insufflation, including procedures performed in the prone or lateral decubitus position. Surgical position was evaluated in subgroup analyses.

### Participants

All adult patients who underwent thoracoscopic esophageal cancer surgery, including robot-assisted procedures, with intrathoracic CO₂ insufflation between January 2022 and December 2024 at Aichi Medical University Hospital and Aichi Cancer Center were eligible for inclusion.

Patients with pre-existing chronic kidney disease not requiring dialysis were eligible for inclusion, and the influence of baseline renal function was evaluated in the analyses.

### Exclusion criteria

Patients were excluded if they met any of the following criteria:

- chronic dialysis
- severe preoperative electrolyte abnormality, defined as a preoperative serum potassium level of 5.0 mEq/L or higher
- intraoperative massive bleeding, defined as blood loss of 2000 mL or more

### Data collection

The following data were collected from medical records.

Patient demographics included age, sex, body mass index, and American Society of Anesthesiologists physical status.

Preoperative variables included estimated glomerular filtration rate, history of diabetes mellitus, hypertension, respiratory dysfunction, cardiac disease, and perioperative use of diuretics, insulin, or β2-agonists.

Procedural variables included surgical approach, anesthesia method, surgical time, anesthesia time, surgical position, and duration in the prone or lateral decubitus position.

Hemodynamic and fluid balance variables included minimum mean arterial pressure during the main thoracic phase, intraoperative urine output, fluid volume, transfusion status, norepinephrine use, and total norepinephrine dose.

Arterial blood gas variables included serum potassium, pH, PaCO₂, HCO₃⁻, base excess, anion gap, and lactate. These variables were collected at the following timepoints:

- **Timepoint 1 (T1), baseline:** after induction of anesthesia and before surgical positioning or incision.
- **Timepoint 2 (T2), intervention:** during the thoracic phase under CO₂ insufflation, at the timepoint of maximal respiratory challenge. If arterial blood gas analysis was performed multiple times during this phase, the sample with the lowest arterial pH was used for the primary analysis.

### Outcomes

The primary outcome was the mean difference in serum potassium concentration between baseline and the intraoperative intervention timepoint under CO₂ insufflation.

Secondary outcomes were designed to explore physiological factors associated with potassium elevation and to assess the robustness of the primary finding. These included changes in arterial blood gas and electrolyte variables, subgroup analyses according to surgical position and institution, sensitivity analysis excluding patients with marked metabolic acidosis, and regression analyses to identify factors associated with the magnitude of serum potassium increase.

The change in serum potassium concentration was defined as ΔK. Changes in other variables, including PaCO₂, pH, lactate, and base excess, were calculated in the same manner.

### Sample size

The sample size rationale was described in the previously published protocol. In brief, preliminary data suggested a clinically relevant increase in serum potassium during the thoracic phase, and a minimum of 34 patients was estimated to provide 80% power to detect a mean paired difference of 0.5 mEq/L with a two-sided alpha level of 0.05.

In the present study, all eligible patients from both institutions during the study period were included to improve precision and allow subgroup and regression analyses.

### Statistical analysis

The selection of the arterial blood gas sample with the lowest pH during the intervention phase was prespecified. This approach was used to focus the analysis on the physiological response, particularly potassium homeostasis, during the most severe degree of acute acidosis encountered during the procedure.

Continuous variables were summarized as mean ± standard deviation or median with interquartile range, as appropriate. Categorical variables were summarized as numbers and percentages.

For the primary outcome, serum potassium concentrations at baseline and the intervention timepoint were compared using a paired t-test.

For secondary analyses, changes in arterial blood gas and electrolyte variables were similarly compared between baseline and the intervention timepoint. Subgroup analyses were performed according to surgical position and institution. A sensitivity analysis was performed after excluding patients with marked metabolic acidosis, defined as base excess < −5 mmol/L at the intervention timepoint.

Univariable linear regression analyses were performed to explore associations between ΔK and candidate physiological or clinical variables, including ΔPaCO₂, ΔpH, lactate at T2, Δbase excess, and preoperative eGFR.

A multivariable linear regression model including ΔPaCO₂, ΔLac, and preoperative eGFR was then constructed. ΔLac, rather than lactate at T2, was included to capture the intraoperative change in metabolic stress from baseline. These variables were selected based on clinical relevance while avoiding overfitting and multicollinearity. Continuous predictors were mean-centered to allow clinically meaningful interpretation of the intercept.

Complete-case analysis was performed for each model, and the number of observations included in each analysis was reported in the corresponding tables and figures.

All statistical analyses were performed using R version 4.5.1 (R Foundation for Statistical Computing, Vienna, Austria). A two-sided p-value of less than 0.05 was considered statistically significant.

### Ethics

This study was centrally reviewed and approved by the Institutional Review Board of Aichi Medical University (approval no. 2025-088). Authorization to conduct the study was subsequently obtained from Aichi Cancer Center. The requirement for individual written informed consent was waived because this was a retrospective study using anonymized data. An opt-out policy was implemented through institutional websites. All data were anonymized before analysis.

The study protocol was registered with the University Hospital Medical Information Network Clinical Trials Registry (UMIN-CTR; UMIN000058829) and published previously [5].

## Results

### Study population

A total of 131 adult patients who underwent thoracoscopic esophagectomy between January 2022 and December 2024 were assessed for eligibility. No patients met any of the predefined exclusion criteria: chronic dialysis, preoperative serum potassium ≥5.0 mEq/L, or massive intraoperative bleeding ≥2000 mL. Paired potassium data at T1 and T2 were available for all patients. Therefore, all 131 patients were included in the final analysis (Fig 1).

**Fig 1.**
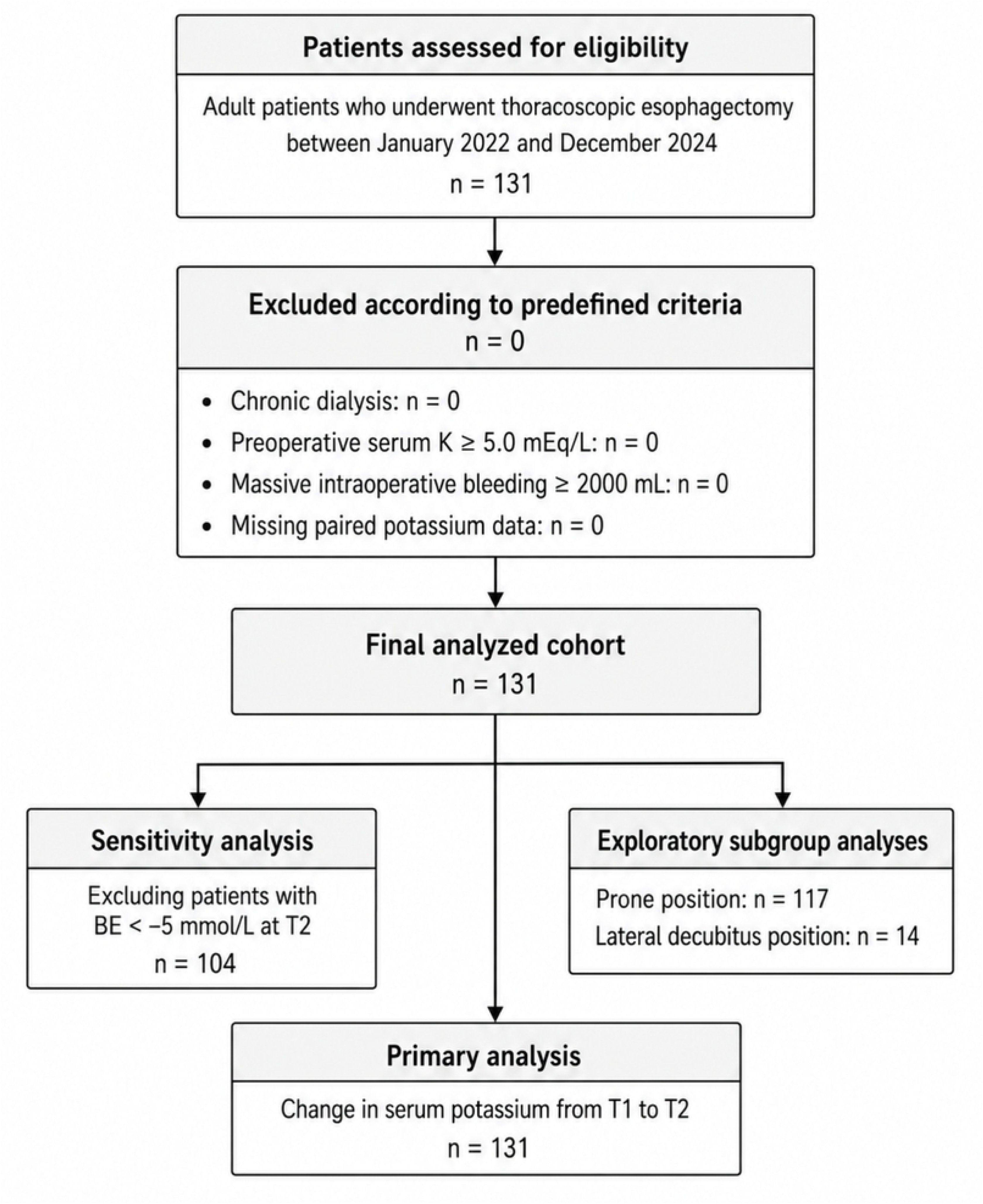
Study flow diagram.

Patient characteristics and perioperative variables are shown in Table 1. The mean age was 68.4 ± 8.9 years, and 106 patients (80.9%) were male. The mean preoperative serum potassium concentration was 4.30 ± 0.40 mEq/L, and the mean preoperative eGFR was 72.6 ± 19.0 mL/min/1.73 m². Most procedures were performed in the prone position (117 patients, 89.3%), while 14 patients (10.7%) underwent surgery in the lateral decubitus position. Robot-assisted procedures were performed in 34 patients (26.0%). The median blood loss was 140 mL [80–240], and norepinephrine was used in 108 patients (82.4%).

**Table 1.** Patient characteristics and perioperative variables.

| Variable | Overall cohort (N = 131) |
| --- | --- |
| **Patient characteristics** |  |
| Age, years | 68.4 ± 8.9 |
| Male sex | 106 (80.9) |
| Body mass index, kg/m<sup>2</sup> | 21.5 ± 3.4 |
| ASA physical status |  |
| 1 | 25 (19.1) |
| 2 | 95 (72.5) |
| 3 | 11 (8.4) |
| Preoperative serum potassium, mEq/L | 4.30 ± 0.40 |
| Preoperative eGFR, mL/min/1.73 m<sup>2</sup> | 72.6 ± 19.0 |
| **Comorbidities** |  |
| Cardiovascular disease | 15 (11.5) |
| Respiratory disease | 14 (10.7) |
| Renal disease | 8 (6.1) |
| **Surgical and anesthetic variables** |  |
| Study site |  |
| Aichi Cancer Center | 106 (80.9) |
| Aichi Medical University Hospital | 25 (19.1) |
| Surgical position |  |
| Prone position | 117 (89.3) |
| Lateral decubitus position | 14 (10.7) |
| Robot-assisted procedure | 34 (26.0) |
| Anesthetic technique |  |
| General anesthesia with epidural anesthesia | 122 (93.1) |
| General anesthesia with regional block | 9 (6.9) |
| Anesthesia time, min | 642 ± 98 |
| Surgical time, min | 555 ± 94 |
| Duration in prone/lateral position, min | 309 ± 86 |
| **Intraoperative management** |  |
| Infusion volume, mL | 3850 [3350–4550] |
| Urine output, mL | 450 [300–648] |
| Blood loss, mL | 140 [80–240] |
| Norepinephrine use | 108 (82.4) |
| Total norepinephrine dose among users, mg | 1.30 [0.73–2.42] |

Data are presented as mean ± SD, median [interquartile range], or n (%).
Duration in prone/lateral position was calculated using available data (n = 130).
Norepinephrine use was defined as cases with a recorded total norepinephrine dose.

### Changes in serum potassium and arterial blood gas variables

Serum potassium significantly increased from 3.96 ± 0.38 mEq/L at T1 to 4.59 ± 0.63 mEq/L at T2. The mean increase in serum potassium was +0.64 mEq/L (95% CI, 0.55 to 0.73; p < 0.001) (Table 2, Fig 2).

**Fig 2.**
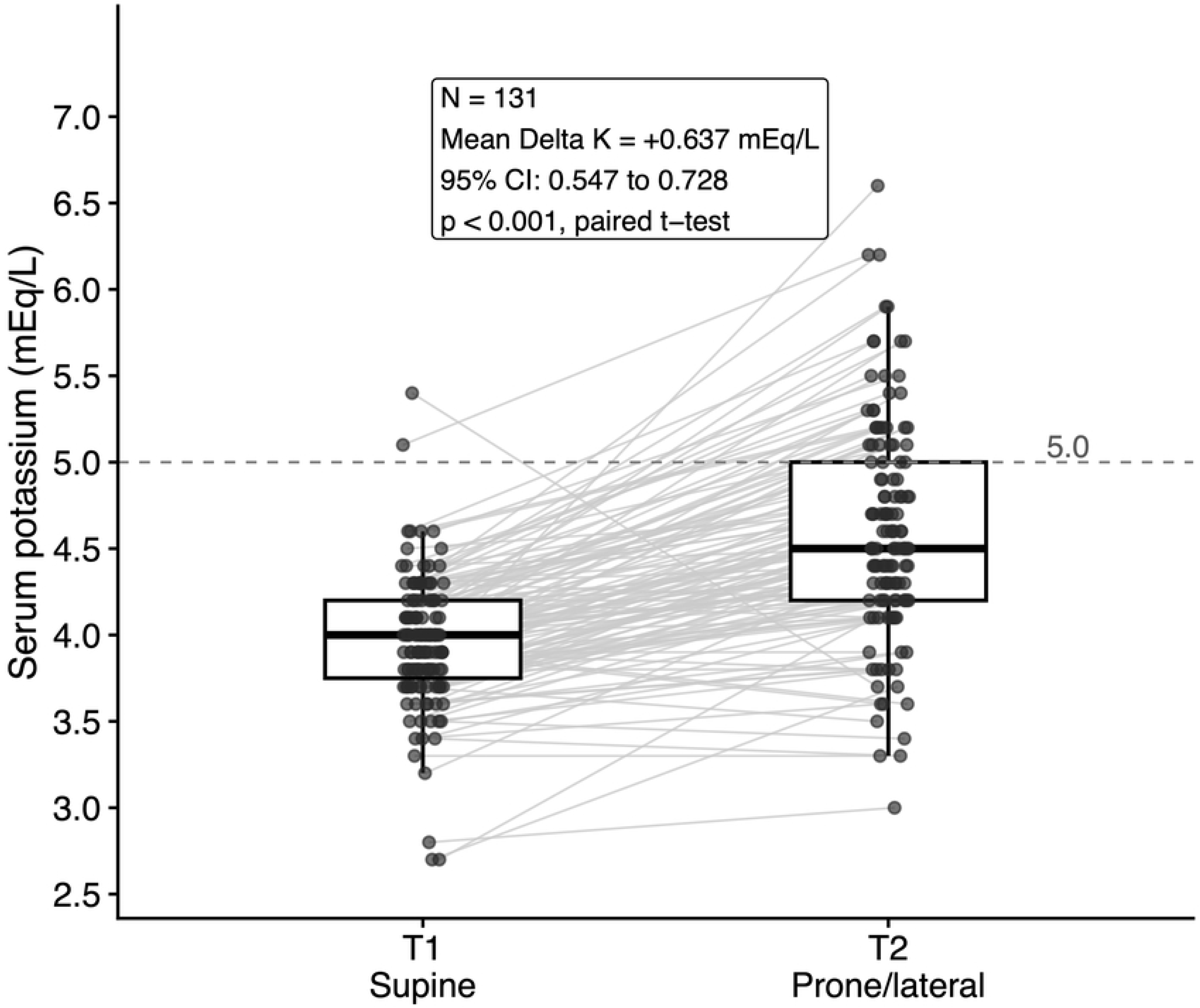
Change in serum potassium concentration from T1 to T2. Serum potassium concentration increased significantly from baseline after anesthesia induction and before positioning (T1) to the intraoperative timepoint after positioning and during intrathoracic carbon dioxide insufflation when respiratory acidosis was most pronounced (T2).

**Table 2.** Changes in arterial blood gas and electrolyte variables.

| Variable | T1 (Supine) | T2 (Prone/lateral) | Mean change (95% CI) | P value |
| --- | --- | --- | --- | --- |
| Serum potassium, mEq/L | $3.96 \pm 0.38$ | $4.59 \pm 0.63$ | $+0.64$ (0.55 to 0.73) | $<0.001$ |
| pH | $7.387 \pm 0.046$ | $7.247 \pm 0.106$ | $-0.140$ (-0.160 to -0.121) | $<0.001$ |
| PaCO<sub>2</sub>, mmHg | $41.48 \pm 5.41$ | $58.60 \pm 18.52$ | $+17.13$ (13.82 to 20.43) | $<0.001$ |
| HCO<sub>3</sub><sup>-</sup>, mmol/L | $24.37 \pm 2.49$ | $23.81 \pm 3.43$ | $-0.56$ (-1.14 to 0.02) | 0.058 |
| Base excess, mmol/L | $-0.58 \pm 3.35$ | $-3.34 \pm 2.48$ | $-2.75$ (-3.28 to -2.22) | $<0.001$ |
| Lactate, mmol/L | $1.08 \pm 0.25$ | $1.09 \pm 0.44$ | $+0.00$ (-0.07 to 0.08) | 0.942 |
| Anion gap, mmol/L | $12.41 \pm 2.98$ | $11.58 \pm 2.66$ | $-0.83$ (-1.52 to -0.14) | 0.019 |

Data are presented as mean ± SD or mean change (95% confidence interval).
P values were calculated using paired t-tests.
Base excess was available for 130 paired measurements; anion gap was available for 106 paired measurements. Other variables were available for 131 paired measurements.

During the same intraoperative period, marked respiratory acidosis was also observed. The mean pH decreased from 7.387 ± 0.046 to 7.247 ± 0.106, with a mean change of −0.140 (95% CI, −0.160 to −0.121; p < 0.001). PaCO₂ increased from 41.48 ± 5.41 mmHg to 58.60 ± 18.52 mmHg, with a mean change of +17.13 mmHg (95% CI, 13.82 to 20.43; p < 0.001). Base excess also decreased significantly, whereas lactate did not change significantly.

### Subgroup and sensitivity analyses

The increase in serum potassium was consistently observed in subgroup and sensitivity analyses (Fig 3). At Aichi Cancer Center, the mean ΔK was +0.637 mEq/L (95% CI, 0.530 to 0.743; p < 0.001; n = 106). At Aichi Medical University Hospital, the mean ΔK was +0.640 mEq/L (95% CI, 0.474 to 0.806; p < 0.001; n = 25).

**Fig 3.**
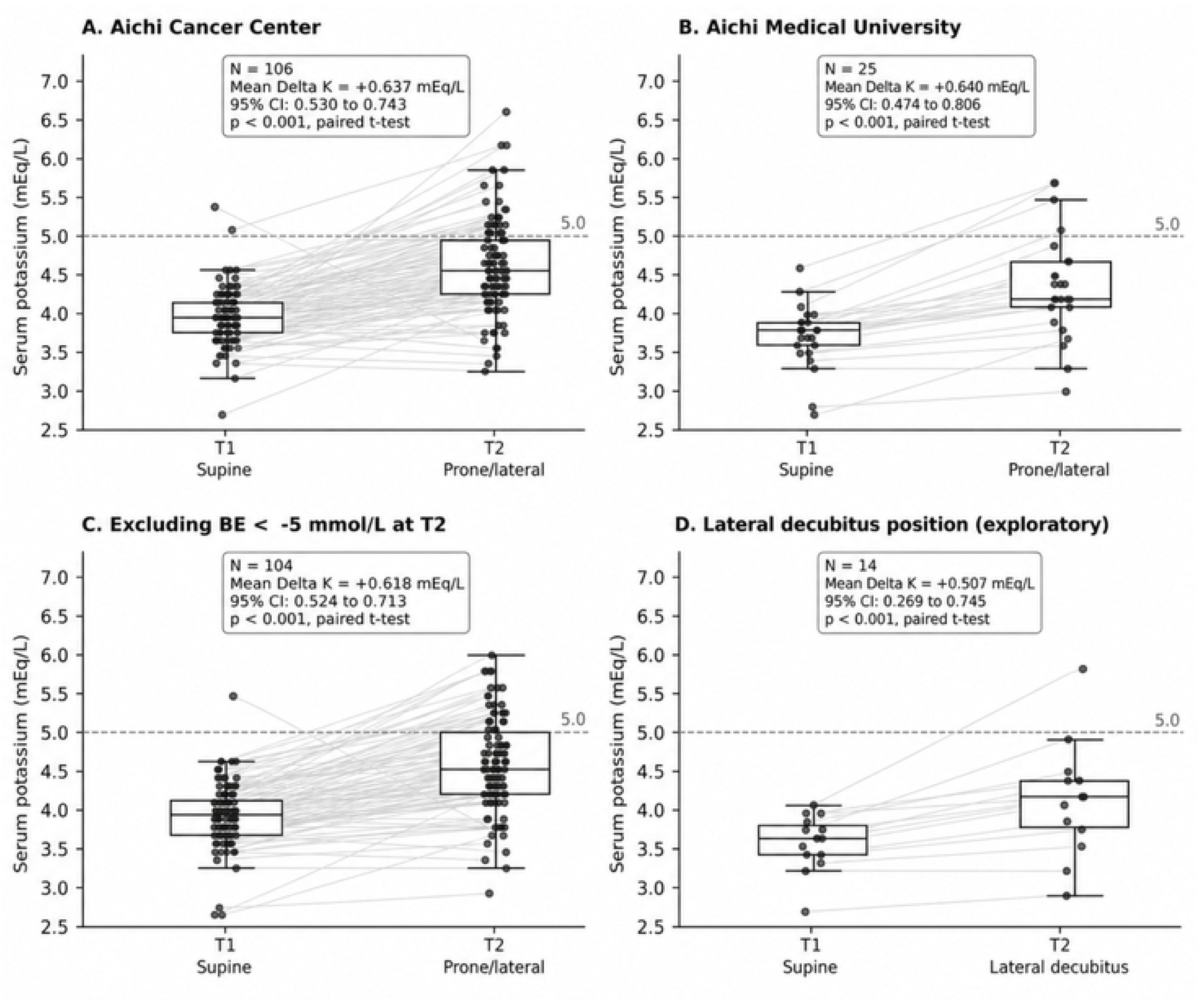
Subgroup and sensitivity analyses of serum potassium change. Changes in serum potassium concentration from T1 to T2 are shown according to study center, patient position, and the sensitivity analysis excluding patients with marked metabolic acidosis, defined as base excess < -5 mmol/L at T2.

After excluding patients with marked metabolic acidosis, defined as base excess < −5 mmol/L at T2, the potassium increase remained significant, with a mean ΔK of +0.618 mEq/L (95% CI, 0.524 to 0.713; p < 0.001; n = 104). In the exploratory analysis limited to patients in the lateral decubitus position, serum potassium also increased significantly, with a mean ΔK of +0.507 mEq/L (95% CI, 0.269 to 0.745; p < 0.001; n = 14).

### Factors associated with potassium increase

Univariable linear regression analyses for ΔK are shown in Table 3. Male sex, longer duration in the prone or lateral position, longer surgical time, higher total norepinephrine dose, and higher lactate at T2 were associated with greater ΔK. In contrast, preoperative eGFR, baseline serum potassium at T1, study site, robot-assisted procedure, lateral decubitus position, ΔPaCO₂, pH decrease, and base excess decrease were not significantly associated with ΔK.

**Table 3.**
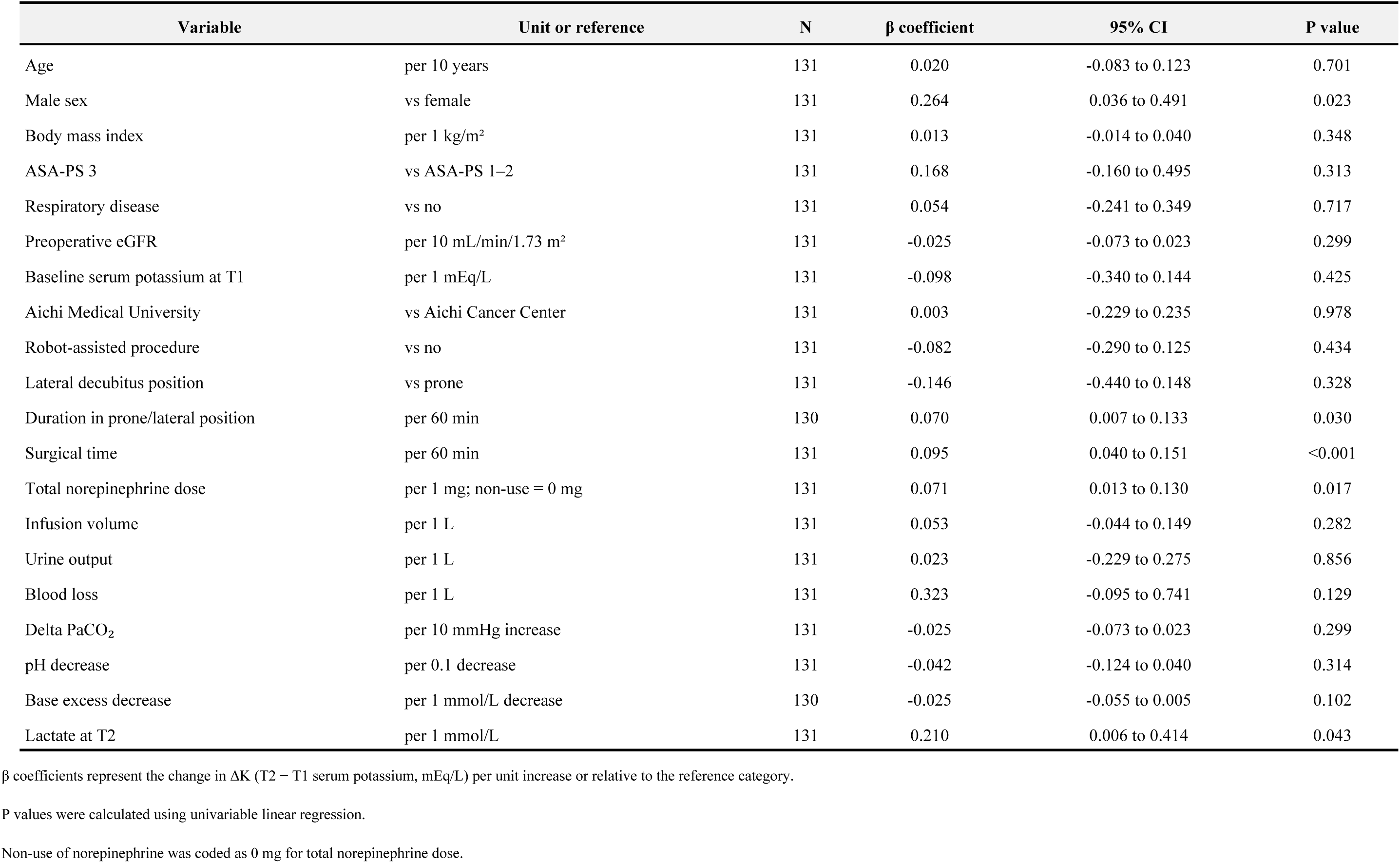
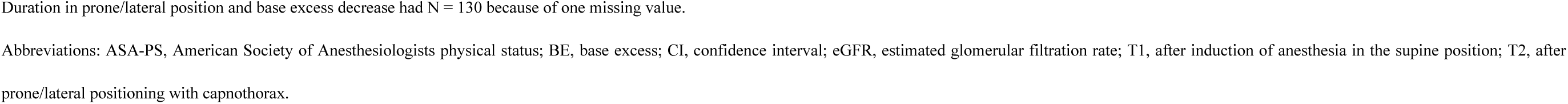
Univariable linear regression analyses for ΔK.

| Variable | Unit or reference | N | $\beta$ coefficient | 95% CI | P value |
| --- | --- | --- | --- | --- | --- |
| Age | per 10 years | 131 | 0.020 | -0.083 to 0.123 | 0.701 |
| Male sex | vs female | 131 | 0.264 | 0.036 to 0.491 | 0.023 |
| Body mass index | per 1 kg/m<sup>2</sup> | 131 | 0.013 | -0.014 to 0.040 | 0.348 |
| ASA-PS 3 | vs ASA-PS 1–2 | 131 | 0.168 | -0.160 to 0.495 | 0.313 |
| Respiratory disease | vs no | 131 | 0.054 | -0.241 to 0.349 | 0.717 |
| Preoperative eGFR | per 10 mL/min/1.73 m<sup>2</sup> | 131 | -0.025 | -0.073 to 0.023 | 0.299 |
| Baseline serum potassium at T1 | per 1 mEq/L | 131 | -0.098 | -0.340 to 0.144 | 0.425 |
| Aichi Medical University | vs Aichi Cancer Center | 131 | 0.003 | -0.229 to 0.235 | 0.978 |
| Robot-assisted procedure | vs no | 131 | -0.082 | -0.290 to 0.125 | 0.434 |
| Lateral decubitus position | vs prone | 131 | -0.146 | -0.440 to 0.148 | 0.328 |
| Duration in prone/lateral position | per 60 min | 130 | 0.070 | 0.007 to 0.133 | 0.030 |
| Surgical time | per 60 min | 131 | 0.095 | 0.040 to 0.151 | <0.001 |
| Total norepinephrine dose | per 1 mg; non-use = 0 mg | 131 | 0.071 | 0.013 to 0.130 | 0.017 |
| Infusion volume | per 1 L | 131 | 0.053 | -0.044 to 0.149 | 0.282 |
| Urine output | per 1 L | 131 | 0.023 | -0.229 to 0.275 | 0.856 |
| Blood loss | per 1 L | 131 | 0.323 | -0.095 to 0.741 | 0.129 |
| Delta PaCO<sub>2</sub> | per 10 mmHg increase | 131 | -0.025 | -0.073 to 0.023 | 0.299 |
| pH decrease | per 0.1 decrease | 131 | -0.042 | -0.124 to 0.040 | 0.314 |
| Base excess decrease | per 1 mmol/L decrease | 130 | -0.025 | -0.055 to 0.005 | 0.102 |
| Lactate at T2 | per 1 mmol/L | 131 | 0.210 | 0.006 to 0.414 | 0.043 |

$\beta$ coefficients represent the change in $\Delta K$ (T2 – T1 serum potassium, mEq/L) per unit increase or relative to the reference category.
P values were calculated using univariable linear regression.
Non-use of norepinephrine was coded as 0 mg for total norepinephrine dose.
Duration in prone/lateral position and base excess decrease had N = 130 because of one missing value.
Abbreviations: ASA-PS, American Society of Anesthesiologists physical status; BE, base excess; CI, confidence interval; eGFR, estimated glomerular filtration rate; T1, after induction of anesthesia in the supine position; T2, after
prone/lateral positioning with capnothorax.

In the centered multivariable linear regression model, ΔLac was independently associated with ΔK (β = 0.303; 95% CI, 0.096 to 0.509; p = 0.004) (Fig 4). In contrast, ΔPaCO₂ (β = −0.004 per 10 mmHg; 95% CI, −0.053 to 0.044; p = 0.860) and preoperative eGFR (β = −0.029 per 10 mL/min/1.73 m²; 95% CI, −0.076 to 0.019; p = 0.236) were not independently associated with ΔK. The intercept remained significantly positive (β = 0.637; 95% CI, 0.549 to 0.726; p < 0.001).

**Fig 4.**
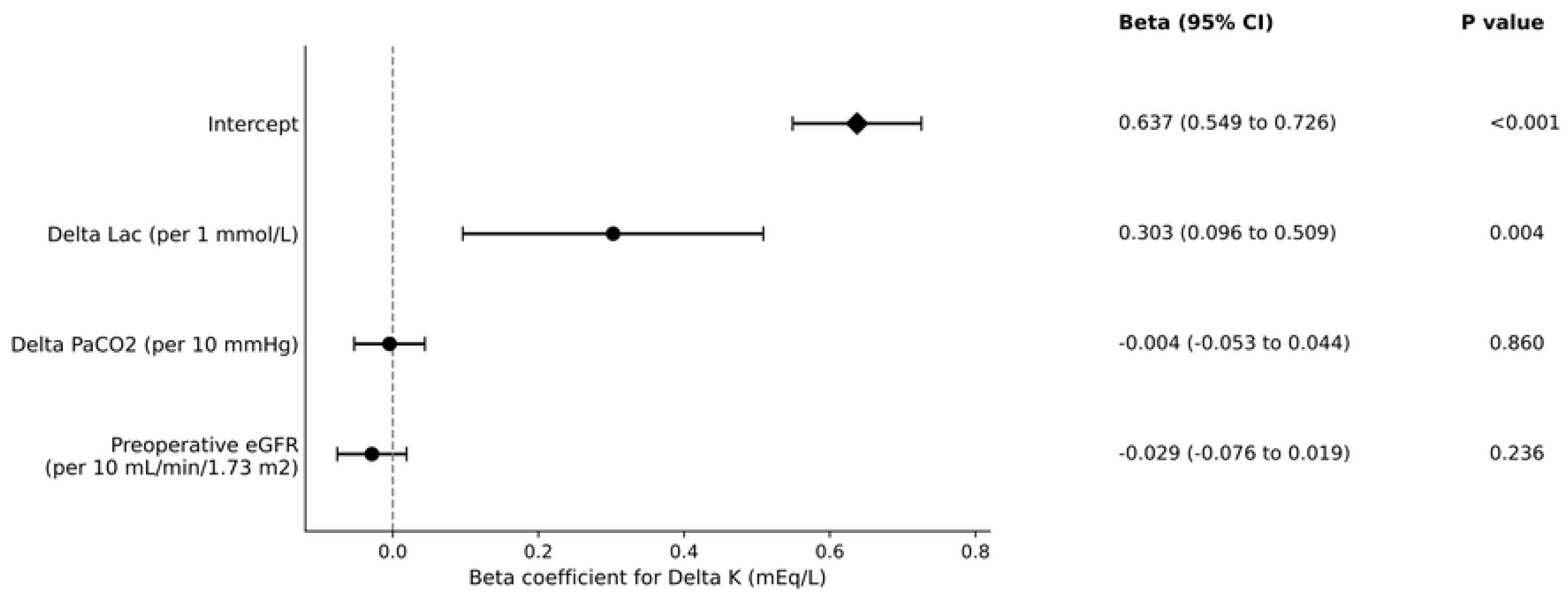
Multivariable linear regression analysis for serum potassium change. Regression coefficients and 95% confidence intervals are shown for the centered multivariable model including change in arterial carbon dioxide tension per 10 mmHg, change in lactate concentration per 1 mmol/L, and preoperative estimated glomerular filtration rate per 10 mL/min/1.73 m².

Detailed results of the univariable and multivariable linear regression analyses are provided in the Supporting Information (S1 Table).

## Discussion

In this analysis, serum potassium increased clearly from baseline to the intraoperative intervention timepoint during thoracoscopic esophageal cancer surgery with intrathoracic CO₂ insufflation. The mean increase was approximately 0.64 mEq/L (Table 2, Fig 2), which was not only statistically significant but also clinically relevant in perioperative management [6]. During the same intraoperative period, acute respiratory acidosis, characterized by a decrease in pH and an increase in PaCO₂, was also observed. Although respiratory acidosis has generally been considered to cause only minimal potassium elevation compared with metabolic acidosis [2, 3], our findings indicate that a more distinct potassium shift may be observed in the specific clinical setting of thoracoscopic CO₂ insufflation.

This finding has direct implications for perioperative management during thoracoscopic esophageal cancer surgery. In settings where CO₂ insufflation, one-lung ventilation, partial lung collapse, and positional changes coexist, PaCO₂ elevation and serum potassium changes may progress even when SpO₂ is preserved. In particular, the PaCO₂–EtCO₂ gradient may widen during this procedure, and EtCO₂ alone may underestimate the severity of acute respiratory acidosis [7]. Therefore, when significant hypercapnia is suspected, arterial blood gas analysis should be used to assess PaCO₂, pH, and serum potassium. In addition, thoracic manipulation is performed near the heart, creating a clinical setting in which intraoperative arrhythmias may occur [8].

Acute serum potassium elevation may amplify this arrhythmic risk, making early recognition of electrolyte changes important for perioperative safety [6, 9].

From a mechanistic perspective, ΔLac was independently associated with ΔK in the multivariable analysis, suggesting that metabolic stress related to surgical stress or circulatory changes may have contributed to the magnitude of potassium elevation [2]. However, the potassium increase was robust to the exclusion of patients with BE < −5 mmol/L, remaining statistically significant and similar in magnitude. This finding indicates that marked metabolic acidosis alone is unlikely to explain the observed increase. Furthermore, the significantly positive intercept in the centered multivariable model (Fig 4) indicates that a substantial potassium increase was predicted even for a patient with average values of ΔLac, ΔPaCO₂, and preoperative renal function. Thus, although ΔLac may explain some of the interindividual variation in ΔK, its association does not establish that metabolic stress alone accounts for the overall potassium increase. The intercept does not identify the underlying mechanism, but it leaves open the possibility that physiological factors shared across the surgical procedure, including hyperacute exogenous CO₂ loading, contributed to the observed increase.

Taken together, these findings cannot be fully interpreted as a simple reflection of metabolic stress. Rather, they suggest that unexpectedly large potassium elevation may occur in a specific intraoperative environment involving intrathoracic CO₂ insufflation, one-lung ventilation, lung collapse, positional change, and hyperacute exogenous CO₂ loading. The fact that ΔPaCO₂ was not an independent predictor does not necessarily negate the contribution of PaCO₂ elevation itself. Instead, it may indicate that a single measured change in PaCO₂ cannot fully capture this complex intraoperative physiological process. In hyperacute CO₂ loading, immediate transcellular shifts, including intracellular H⁺ buffering and K⁺ movement, may become prominent before slower renal compensatory mechanisms become effective [2, 4]. Such a difference in time scale may partly explain why potassium elevation occurred in a setting where conventional respiratory acidosis would not usually be expected to cause a marked potassium shift [3].

In subgroup analyses, similar potassium increases were observed across both institutions. In the exploratory analysis of patients in the lateral decubitus position, serum potassium also increased significantly. These findings suggest that the potassium increase may not be limited to the prone position itself, but may instead be related to the broader thoracoscopic surgical environment with CO₂ insufflation. However, because the number of lateral decubitus cases was small, this finding should be interpreted as hypothesis-generating.

These findings may also have implications beyond the operating room, particularly for the management of acute hypercapnia in the ICU. In patients with respiratory failure such as ARDS, permissive hypercapnia is sometimes accepted as part of lung-protective ventilation, but the effect of acute CO₂ elevation on electrolyte homeostasis remains insufficiently understood. Moreover, excessive deviation of PaCO₂ has been associated with worse outcomes in patients with acute brain injury [10], whereas moderate permissive hypercapnia during perioperative esophagectomy has been reported to be associated with reduced postoperative delirium [11]. Thus, the acceptable range of hypercapnia may differ according to the underlying condition and clinical context.

Although the present analysis focused on a specific intraoperative setting and cannot be directly extrapolated to ICU patients, it suggests that serum potassium changes deserve attention during hyperacute respiratory acidosis.

Highly specialized surgical and anesthetic procedures can give rise to acute physiological conditions that are rarely encountered or systematically studied in routine clinical care. Hyperacute respiratory acidosis, for example, is difficult to evaluate at comparable levels of severity and at standardized timepoints in ordinary disease states. Thoracoscopic esophagectomy involving intrathoracic CO₂ insufflation, one-lung ventilation, and lung collapse may therefore provide a nonphysiological but informative clinical model for examining physiological responses to acute CO₂ loading. A similar rationale underlay our previous study of anesthesia for airway stenting in patients with severe airway stenosis, in which controlled positive-pressure ventilation with muscle relaxants reduced hypoxic events despite conventional concerns regarding the use of muscle relaxants in these patients [12]. Although the clinical questions differ, both studies illustrate how research in specialized surgical and anesthetic settings can challenge conventional assumptions and reveal clinically relevant physiological responses that would otherwise be difficult to investigate.

This analysis has several strengths. First, the paired design, in which each patient served as their own control, allowed assessment of acute changes while minimizing the influence of interpatient variability. Second, the inclusion of patients from two institutions provides a certain degree of generalizability compared with a single-center study. Third, arterial blood gas data allowed simultaneous evaluation of changes in pH, PaCO₂, BE, lactate, and serum potassium. In addition, the study was conducted according to a previously published protocol, enhancing transparency regarding the prespecified primary outcome and core analytical framework.

Several limitations should also be acknowledged. First, because of the retrospective design, the timing of blood gas sampling and anesthetic management were not fully standardized. Second, although this was a two-center study, most patients were from one institution, resulting in an imbalance in case numbers between centers. Third, the number of lateral decubitus cases was small, limiting comparison of the effect of surgical position. Fourth, postoperative ICU potassium values were not analyzed because sampling times varied substantially and were not directly aligned with the primary focus of this study, namely acute intraoperative changes during CO₂ insufflation. Therefore, this study could not determine how quickly the observed acute potassium elevation returned to baseline after cessation of CO₂ insufflation, or whether it persisted into the early postoperative period. This issue should be addressed in future studies with standardized postoperative sampling timepoints. Finally, in an intraoperative environment where respiratory and metabolic factors coexist, it is difficult to completely separate their respective contributions.

In conclusion, thoracoscopic esophageal cancer surgery with intrathoracic CO₂ insufflation was associated with a clinically meaningful increase in serum potassium; acute respiratory acidosis was also observed during the same intraoperative period. This increase persisted even after excluding patients with marked metabolic acidosis, suggesting that it may not be explained solely by metabolic stress. In this specific intraoperative environment involving CO₂ insufflation, lung collapse, and positional change, attention should be paid to serum potassium fluctuations, and arterial blood gas analysis is important for appropriate assessment.

## Data Availability

The data underlying this study cannot be shared publicly because they contain potentially identifying clinical information and are subject to ethical and privacy restrictions. Data may be made available upon reasonable request to qualified researchers who meet the criteria for access to confidential data, with approval from the Institutional Review Board of Aichi Medical University. Requests may be directed to the Aichi Medical University Institutional Review Board.

## Supporting information

S1 Table. Full results of the univariable and multivariable linear regression analyses.

